# Improved Quantitative Parameter Estimation for Prostate T2 Relaxometry using Convolutional Neural Networks

**DOI:** 10.1101/2023.01.11.23284194

**Authors:** Patrick J. Bolan, Sara L. Saunders, Kendrick Kay, Mitchell Gross, Mehmet Akcakaya, Gregory J. Metzger

## Abstract

This work seeks to evaluate multiple methods for quantitative parameter estimation from standard T_2_ mapping acquisitions in the prostate. The T_2_ estimation performance of methods based on neural networks (NN) was quantitatively compared to that of conventional curve fitting techniques. Large physics-based synthetic datasets simulating T_2_ mapping acquisitions were generated for training NNs and for quantitative performance comparisons. Ten combinations of different NN architectures, training strategies, and training corpora were implemented and compared with four different curve fitting strategies. All methods were compared quantitatively using synthetic data with known ground truth, and further compared on *in vivo* test data, with and without noise augmentation, to evaluate feasibility and noise robustness. In the evaluation on synthetic data, a convolutional neural network (CNN), trained in a supervised fashion using synthetic data generated from naturalistic images, showed the highest overall accuracy and precision amongst all the methods. On *in vivo* data, this best-performing method produced low-noise T_2_ maps and showed the least deterioration with increasing input noise levels. This study showed that a CNN, trained with synthetic data in a supervised manner, may provide superior T_2_ estimation performance compared to conventional curve fitting, especially in low signal-to-noise regions.

## 1. Introduction

Quantitative T_2_ mapping provides a more objective and potentially sensitive imaging biomarker for diagnosis and grading of prostate cancer compared to qualitative T_2_-weighted imaging ^1–4^. Mapping is conventionally performed in a two-step process. The first step generates a series of images with increasing echo times, typically using a multi-echo spin-echo acquisition. The second step performs curve fitting on a pixel-by-pixel basis across the image series to estimate the exponential signal decay constant, T_2_. This two-step processing is common to a larger group of quantitative MR methods, including other relaxometry applications (T_1_, T_2_, T _2_^*^, T_1rho_, etc.), estimation of apparent diffusion coefficient or intravoxel incoherent motion (IVIM) parameters from diffusion-weighted images (DWI), and measurement of flip angle maps for system calibrations. The fitting step requires images with a high signal-to-noise ratio (SNR) and a wide range of echo times to accurately estimate T_2_ values. This need for multiple images and high SNR leads to long acquisition times, which limits the adoption of these potentially valuable measurements in both clinical and research applications.

A variety of approaches have been developed for reducing acquisition times by merging the acquisition and estimation steps. These approaches include magnetic resonance fingerprinting ^5– 7^, model-based inverse reconstructions ^8–11^ or deep-learning methods ^12–21^ that directly estimate parameter maps from undersampled k-space data. These end-to-end reconstruction techniques can greatly reduce acquisition times, but they require specialized acquisition techniques, and cannot be used to retrospectively process images generated by conventional acquisitions.

In this work we focus on the second part of the conventional processing approach, estimating the transverse relaxation time (T_2_) from a series of fully-reconstructed magnitude images obtained from a multi-echo acquisition. Estimation is often performed on magnitude images because they are routinely generated by MR scanners and are readily available for both prospective and retrospective studies. The standard technique for this processing, using pixel-wise iterative non-linear least squares (NLLS) fitting, is known to overestimate T_2_ when SNR is low ^22,23^. This is because magnitude images have noise with a Rician rather than Gaussian distribution. With Rician noise the measured signal does not decay to zero at long TEs but rather reaches a plateau that depends on the noise level ^22,24^. Minimizing the square of the difference between the measured and estimated data does not give a maximum likelihood estimation of the parameters, as it would if the noise were Gaussian distributed. It is possible to address this problem by performing a true maximum likelihood optimization incorporating the Rician distribution ^25–27^, or using approximations that enable the use of efficient least-squares algorithms ^23,28–30^. All these methods require that the per-pixel noise level is known or can be estimated from the images. This can be very difficult if the noise is spatially varying, as is the case with parallel imaging reconstructions, or if there is not a suitable background region in the images.

Neural networks (NNs) can be used as an alternative to the NLLS fitting step. Several groups have shown that NNs can improve robustness to image noise and are more computationally efficient than iterative fitting. One strategy, previously demonstrated for diffusion ^31^ and T_2_ relaxometry ^32^ problems, is to train a one-dimensionally fully-connected neural network using a large, synthesized training dataset that simulates a forward signal model with random parameter values and Rician noise. These methods improved accuracy and reduced variability compared to the conventional NLLS method. Another group has trained 1D NNs using a self-supervised approach ^33,34^, in which estimated parameters were inserted into the signal model to produce an estimated signal, and the mean squared error between measured and estimated signal was used as a data consistency loss function. Despite the different training strategy this also yielded lower variability of parameter estimates compared to the NLLS method.

The above networks performed inference on a per-pixel basis, and thus did not incorporate information about the spatial correlation of pixels. Such spatial information can be used by using a convolutional neural network (CNN) to operate on patches or whole image sets rather than individual pixels. This approach has been combined with the self-supervised training strategy for both IVIM parameter mapping ^35^ and T_2_ ^*^ relaxometry in the brain ^36^, and gave better noise performance than 1D NNs or NLLS fitting. CNNs trained in a supervised fashion have also been demonstrated to improve noise performance over NLLS fitting in a prostate T2 relaxometry application ^37^.

These prior works demonstrate that NNs can give performance improvements over NLLS fitting in quantitative parameter estimation problems. However, it is not yet clear how these methods produce these benefits. Possible contributing factors include a more accurate representation of Rician noise, denoising capabilities induced by training on noisy inputs, and incorporation of spatial prior information through trained convolutional layers. Improving our understanding of these mechanisms can help researchers recognize the strengths and limitations of these relatively new analysis techniques and use them to provide better quantitative imaging performance.

In this work we expand on these prior developments by implementing multiple NN approaches and systematically comparing their quantitative performance for T_2_ mapping in the prostate. We propose a novel method for synthesizing large datasets suitable for training and testing networks built from a photographic database. Two synthetic test datasets, one with and one without spatial correlation, are used to isolate the contribution of learned spatial priors to method performance. Networks using 1D and convolutional architectures are implemented and trained using both supervised and self-supervised strategies. The performance of these networks, along with several conventional fitting methods, is evaluated on synthetic test datasets with known ground truth, to allow quantitative assessments of bias and precision. Our evaluations focus on performance in low SNR regions, as improved estimation with low SNR data can be used to increase spatial resolution and shorten scan times with higher acceleration factors. Finally, methods are compared on *in vivo* test data, with and without noise augmentation, to evaluate feasibility and noise robustness.

## 2. Methods

### 2.1 Signal Model

In this manuscript we consider only mono-exponential signal decay using normalized parameters. The signal is described as

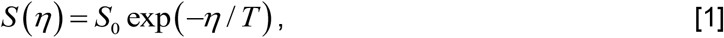

where *η* is the normalized sampling dimension, *T* is the time constant, and *S*_*0*_ is the signal at *η=0*. The sampling dimension is normalized so that the maximum value is 1.0 for a given dataset. This normalization generalizes the problem so that it can describe different sampling times and other mono-exponential processes (e.g., diffusion). For the specific case of T_2_ mapping, the echo time TE and the relaxation time constant T_2_ are both normalized by the maximum echo time: *η*=*TE/TE*_*max*_ and *T =T*_*2*_*/TE*_*max*_.

### 2.2 In Vivo MRI Datasets

All prostate MR imaging was acquired on a Siemens 3T Prisma scanner with surface and endorectal receive coils under an IRB-approved protocol. Multi-echo multi-slice fast spin-echo MR images were acquired from 118 participants using the vendor’s spin-echo multi-contrast sequence, with parameters TR = 6000 ms, eleven echoes with TE = 13.2-145.2 ms in 13.2 ms increments, 256 × 256 images with resolution 1.1 × 1.1 mm, 19-28 axial slices 3 mm thick, accelerated with GRAPPA R=3, total acquisition time 6.5 - 8.5 min. Using the normalized signal model of equation 1, *η* takes values of 0.091, 0.182, 0.272, …, 1.0. An example image series is shown in Figure 1.

**Figure 1.**
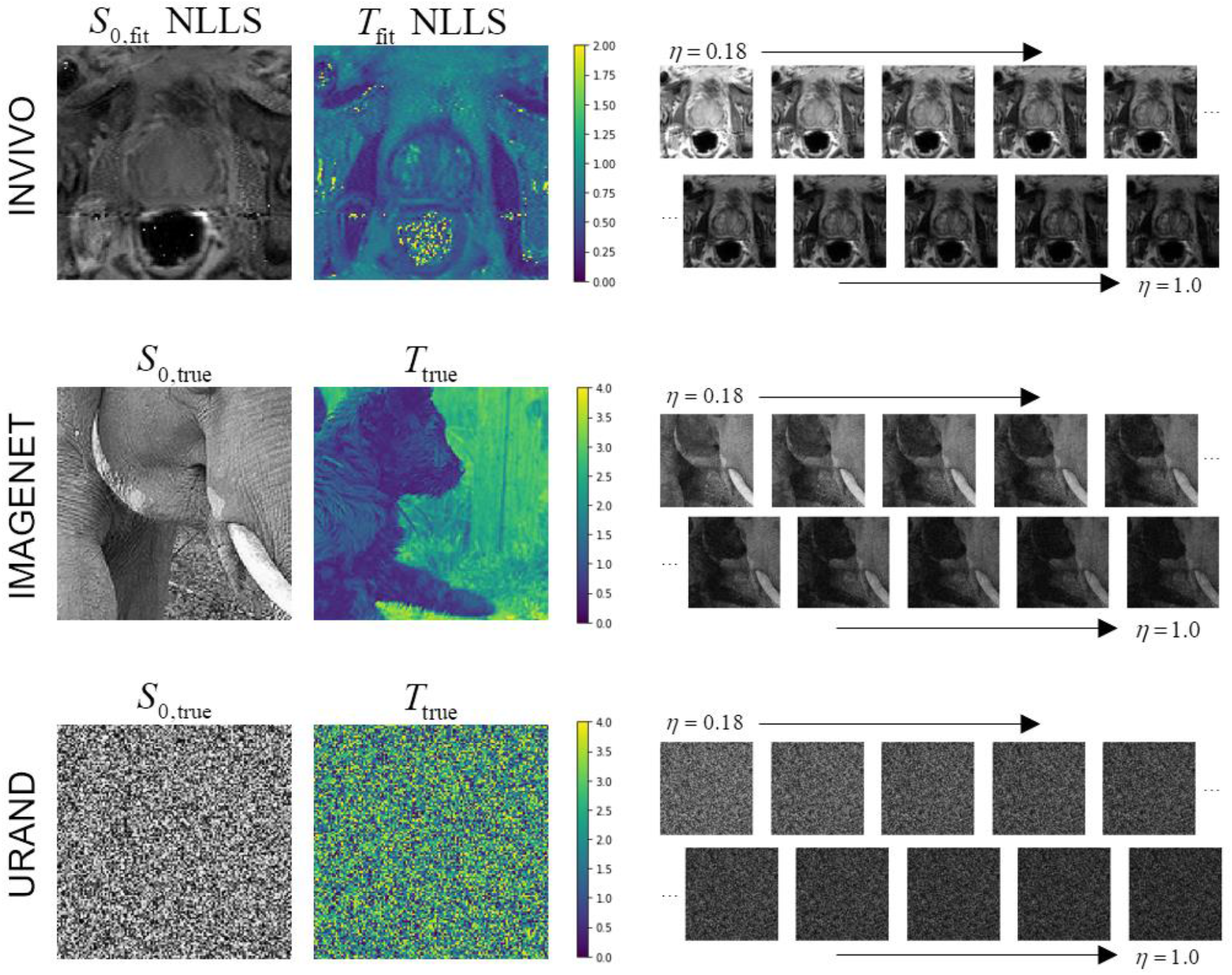
Examples of the three datasets used in the study. The INVIVO data included *T*_2_-weighted images at ten echo points from TE = 26.4 - 145.2 ms, normalized to *η* = 0.18 to 1.0. The *S*_*0*_ and *T* maps from conventional NLLS fitting are shown for this example since the true values are unknown. The IMAGENET dataset used photographic images from the ImageNet collection ^39,40^ as gold-standard *S*_*0*_ and *T*, and synthesized exponential image series, with added Rician noise, matching the *η* values from the INVIVO data. The URAND dataset used a uniform random image for both *S*_*0*_ and *T*, and synthesized an exponential image series in the same manner as for IMAGENET.

Each axial slice from these acquisitions was treated as an independent 3D image series, consisting of two spatial dimensions and one *η* dimension. The intensity of each image series was normalized so that the maximum value over all three dimensions was 1.0. Prior to fitting and analyses, each image series was spatially center-cropped to 128×128 pixels, and the first echo was discarded to avoid stimulated echo contamination ^38^. The full dataset, termed *INVIVO* herein, was randomly split into a test dataset (32 subjects with 695 image series) and a separate training dataset (86 subjects with 1988 image series) used for training NNs.

### 2.3 Synthetic Datasets

Two datasets of simulated T_2_ relaxometry measurements with known ground truth were generated for training and evaluation. The first dataset, called *IMAGENET* herein, was synthesized from a physics-based signal model using naturalistic images drawn from the publicly-available ImageNet dataset ^39,40^ as the gold standard values for *S*_*0*_ and *T*. Using naturalistic images as surrogates for MR images enables the generation of very large datasets, provides a variety of structures and textures, and has been used successfully in training CNNs for MR image reconstruction ^41^. For each generated image series, two unique images were selected for *S*_*0*_ and *T*, converted to floating-point grayscale images, and center-cropped to 128×128 pixels. These were scaled so that *S*_*0*_ ϵ[0,1] and *T* ϵ[0.045, 4], and used to create a series of images *S(η)* from *η* = 0.182, 0.272, …, 1.0 following the signal model of equation 1 and matching the *in vivo* acquisition. Complex Gaussian noise was added to each image, with zero mean and standard deviation *σ* drawn from a uniform random distribution in [0.001, 0.1] for each image series, followed by a magnitude operation. These synthesized images series simulate relaxometry measurements with variable levels of Rician noise, broad parameter ranges, and known ground truth.

The second synthetic dataset, called *URAND*, used random pixel values for the reference images *S*_*0*_ and *T*. In this dataset, spatially adjacent pixels were randomly drawn, so networks trained on this data could not use neighboring pixels to improve accuracy. This approach was designed as a comparison to the *IMAGENET* approach, with the expectation that networks trained on this data would be less dependent on the statistics of training dataset and less prone to blurring artifacts. This dataset was produced in the same manner as the *IMAGENET* dataset but using gold standard *S*_*0*_ and *T* images consisting of random pixel values drawn from a uniform distribution over the same ranges.

Examples of image series from all three datasets are given in Figure 1. For both synthetic datasets, 10,000 image series were generated for *training*, and an additional 1000 image series (with unique *S*_*0*_ and *T* images and noise) were generated to create independent *test* datasets. Figure 2 provides a schema showing the structure of all three datasets.

**Figure 2.**
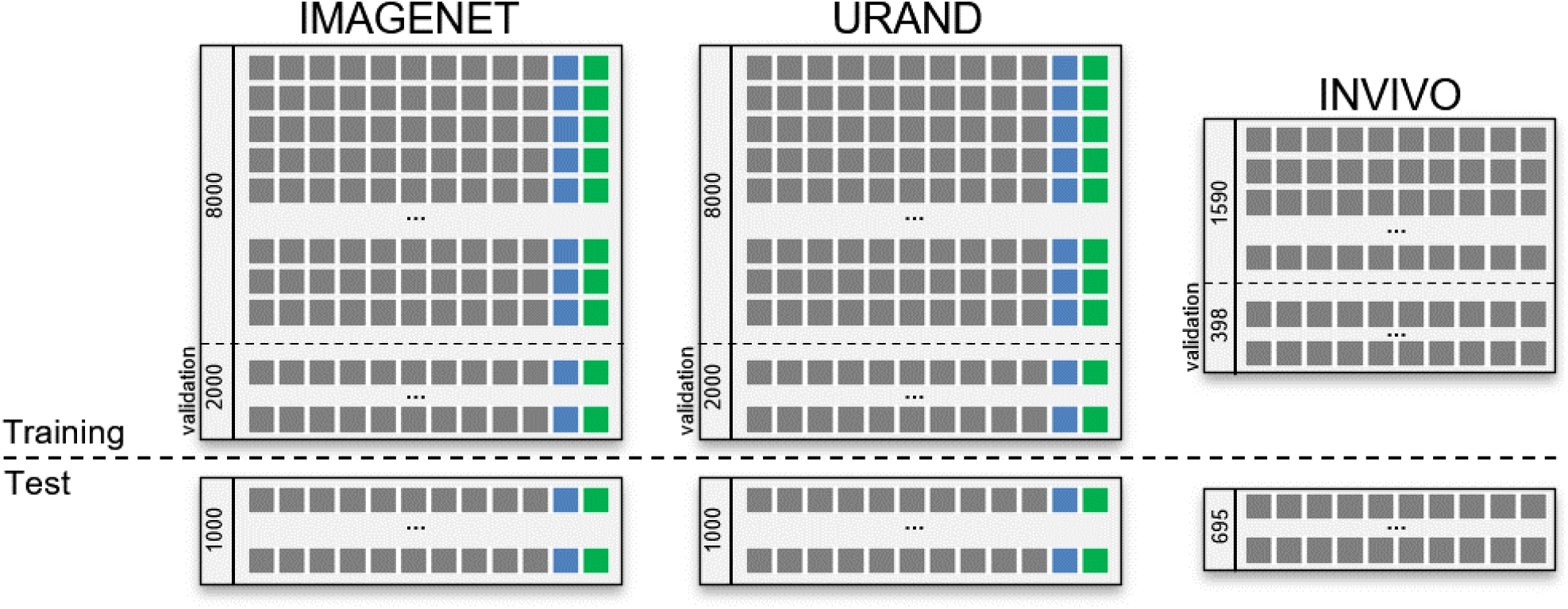
Schema of the datasets used in this study. The upper left box shows the IMAGENET training dataset, consisting of 10,000 series, each including 10 images (gray boxes) with progressively increasing *η* values, and the true *S*_0_ (blue box) and *T*_2_ (green box) maps. Twenty percent of this dataset was reserved for validation during the training process. A separate IMAGENET test dataset of 1000 image series and associated *S*_0_, *T*_2_ labels is used for independent evaluation. The URAND dataset has the same structure, with randomly generated images as described in the text. The INVIVO training and test datasets are smaller than the synthetic datasets and do not include true *S*_0_ and *T*_2_ maps.

### 2.4 Curve Fitting Methods

Four variations of curve fitting were evaluated, which are representative of common practices in quantitative MRI. The FIT_LOGLIN method fit a straight line to the natural logarithm of *S(η)* using the *linalg*.*lstsq()* algorithm in NumPy ^42^. This non-iterative linearized method is widely used due to its speed, but the log transformation effectively increases the weighting on the points with higher signal ^43^. The results from the FIT_LOGLIN method were used as initial guesses for all other methods.

The FIT_NLLS method used the iterative *optimize*.*curve_fit()* method of SciPy ^44^ with the Levenburg-Marquardt algorithm to estimate *S*_*0*_ and *R=1/T* by minimizing the least-squares residuals using equation 1. Note *R* was fit rather than *T* to avoid division-by-zero numerical errors. The FIT_NLLS_BOUND method was similar but used the Trust Region Reflective algorithm with bounds *S*_*0*_ ϵ[0,1000] and *1/T* ϵ[0.25, 22], equivalent to *T* ϵ[0.045, 4], to restrict parameter estimates to physically reasonable values. These two methods, FIT_NLLS and FIT_NLLS_BOUND, assumed a Gaussian noise distribution.

We did not use a true maximum likelihood method with a Rician distribution in this work. This is because our dataset had spatially varying noise (due to parallel imaging), and in our initial evaluations we found that estimating three parameters (*S*_*0*_, *R, σ*_*Rice*_) on a per-pixel basis was numerically unstable, likely due to the Bessel functions that describe the Rician distribution. Therefore we implemented an approximate method that minimized the residual between the measured data and the expectation value of a Rician distribution, as reported by other groups ^23,30^. This method, termed FIT_NLLS_RICE, used *optimize*.*least_squares()* to fit three parameters (*S*_*0*_, *R, σ*_*Rice*_), and used the same algorithm and bounds as FIT_NLLS_BOUND. Table 1 summarizes the four fitting methods used in this work.

**Table 1.** Summary of the four curve fitting methods used in this study. TRF=Trust region reflective, LM=Levenburg-Marquardt

| Method name | Algorithm | Estimated Parameters | Bound Parameters |
| --- | --- | --- | --- |
| FIT_LOGLIN | Linear regression | $S_0, R$ | none |
| FIT_NLLS | LM, iterative | $S_0, R$ | none |
| FIT_NLLS_BOUND | TRF, iterative | $S_0, R$ | $S_0, R$ |
| FIT_NLLS_RICE | TRF, iterative | $S_0, R, \sigma_{\text{Rician}}$ | $S_0, R$ |

### 2.5 Neural Networks

Two neural network architectures were used in this study. The first was a one-dimensional fully-connected neural network used for estimating parameters one pixel at a time. The network had 10 inputs (one for each *η*), 6 hidden layers with 64 weights each, 2 output channels, and used ReLU activations in all layers.

The second network was a 2D convolutional neural network based on the enhanced U-Net provided in the MONAI ^45^ library, which extends the original U-Net ^46^ with residual units in the first 2 downsampling layers ^47^. The network had inputs of 128×128 with 10 input channels (one for each *η*), 4 layers of encoding and decoding with increasing widths [128, 128, 256, 512], 3×3 convolutions in all layers, outputs of 128×128 with two channels (interpreted as *S*_*0*_, *T*), and used batch normalization and PReLU activations.

Both supervised and unsupervised training strategies were used. Supervised training was performed using a mean squared error loss relative to the ground truth *S*_*0*_ and *T*. Self-supervised training was performed as in previous works ^33–36^: the *S*_*0*_ and *T* values produced by the networks were used to simulate values of *S(η)*, and the mean square error between simulated and input data was used as the loss function; thus no ground truth labels were required for the self-supervised training. Note that this self-supervised training approach does not fully account for the Rician distribution of the noise: the estimated signal approaches zero at long *η* values rather than a non-zero value expected with Rician noise. This mismatch is expected to lead to overestimation of *T*, similar to methods that assume Gaussian noise (e.g., FIT_NLLS).

Training the 1D networks was performed by loading each image series and iterating over all spatial positions to extract single-pixel decay curves. Training was performed for 5 epochs using a batch size of 10,000 and the AdamW optimizer ^48^ with learning rate of 0.002. The training sets used 800 image series (thus 128*128*800 = 13.1E6 1D series) for training and 200 image series (3.3E6 1D series) for validation. Convolutional networks were trained for 1000 epochs with a batch size of 100 image series and an AdamW optimizer with learning rate = 0.002. Each training set was further divided into train (80%) and validation (20%) subsets for monitoring training progress.

Combining the two training strategies and three datasets, there were five models trained for each of the two network architectures, producing a total of ten trained NN models for subsequent evaluation. Table 2 summarizes the naming scheme used for all fourteen parameter estimation methods compared in this work, including the 10 trained NN models and the four curve fitting techniques.

**Table 2.**
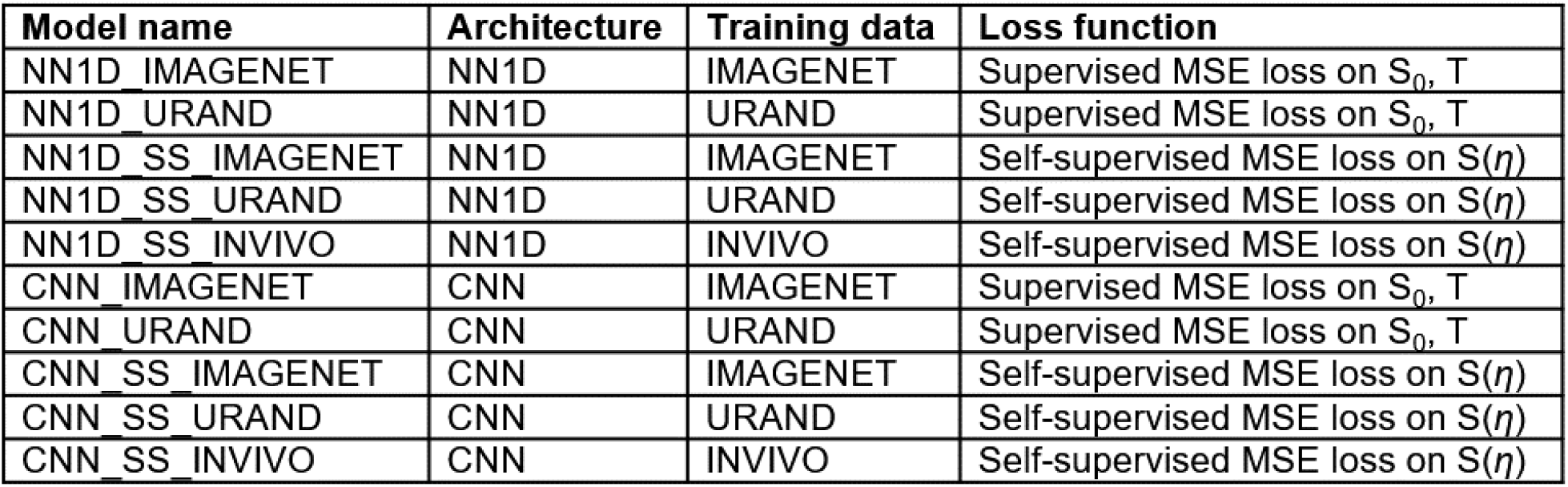
Summary of the ten NN models trained and used in this study

| Model name | Architecture | Training data | Loss function |
| --- | --- | --- | --- |
| NN1D_IMAGENET | NN1D | IMAGENET | Supervised MSE loss on $S_0, T$ |
| NN1D_URAND | NN1D | URAND | Supervised MSE loss on $S_0, T$ |
| NN1D_SS_IMAGENET | NN1D | IMAGENET | Self-supervised MSE loss on $S(\eta)$ |
| NN1D_SS_URAND | NN1D | URAND | Self-supervised MSE loss on $S(\eta)$ |
| NN1D_SS_INVIVO | NN1D | INVIVO | Self-supervised MSE loss on $S(\eta)$ |
| CNN_IMAGENET | CNN | IMAGENET | Supervised MSE loss on $S_0, T$ |
| CNN_URAND | CNN | URAND | Supervised MSE loss on $S_0, T$ |
| CNN_SS_IMAGENET | CNN | IMAGENET | Self-supervised MSE loss on $S(\eta)$ |
| CNN_SS_URAND | CNN | URAND | Self-supervised MSE loss on $S(\eta)$ |
| CNN_SS_INVIVO | CNN | INVIVO | Self-supervised MSE loss on $S(\eta)$ |

### 2.6 Evaluation on Synthetic Data

Comparisons between all 14 parameter estimation methods were performed by evaluating all methods on the two synthetic test datasets (IMAGENET and URAND), which allowed for analysis of error because the ground truth *T* maps are available. Each method was used to estimate a *T* map for each image series in the test datasets. The signed error between true and estimated maps *(T*_pred_*-T*_true_), and the absolute error (|*T*_pred_*-T*_true_|), were calculated for each pixel. Errors were summarized on a per-slice basis by taking the median error value over each map. The median value of the signed error was interpreted as the **bias** of a method. The interquartile range (IQR, 75^th^-25^th^ percentile) of the signed error was interpreted as a measure of **precision**. The median value of the absolute error was interpreted as a measure of overall **accuracy**, which is dependent on both bias and precision, with smaller errors indicating higher accuracy. Decomposing accuracy into separate contributions of bias and precision is important for understand potential sources of bias in quantitative analyses ^49,50^.

Errors were also evaluated on a per-pixel basis to assess their dependence on SNR and *T*. SNR was calculated on a per-pixel basis using

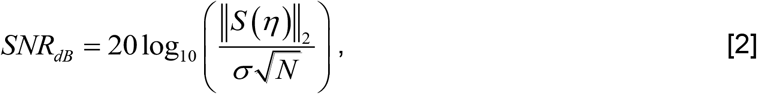

where *σ* is the standard deviation of noise, N is the number of *η* values, and ||…||_2_ is the L2-norm. With this definition the SNR of the synthetic data varied spatial and ranged from negative infinity (where *S*_*0*_ = 0) to 59 dB (for *S*_*0*_ = 1, *T* = 4, *σ* = 0.1), covering a realistic range of *in vivo* values. Finally, each predicted *T* map was compared to the true *T* map using the structural similarity index measure (SSIM) ^51^, a quantitative measure of perceptual similarity between two images, to evaluate the suitability of the method for producing subjectively interpretable *T* maps.

### 2.7 Evaluation on In Vivo Data

The best-performing methods from the synthetic evaluation were subsequently evaluated on the INVIVO test dataset. We compared the results of each method to the results of the FIT_NLLS method because there is no true *T* map available, and FIT_NLLS represents the most conventional approach. The *T* maps were compared qualitatively to determine if their estimation performance was consistent with the findings in the synthetic experiment.

Finally, a noise-addition experiment was performed to evaluate the sensitivity of each method to progressively increasing noise. Complex Gaussian noise with standard deviation ranging from σ = 0.02 to 0.08 units was added to each normalized image series in the INVIVO test dataset, followed by a magnitude operation to generate an image series with increased Rician noise. Original and noise-augmented datasets were used as input to estimate *T* maps using all the methods, without retraining any NNs. Each map was compared to the *T* map generated from the original (no added noise) image series using the same method, so that the per-slice error was the median over *T*_added_noise_ *-T*_original_. Bias, precision, and accuracy with increasing noise levels were interpreted in the same manner as in the synthetic evaluation.

All computation for this work was performed in Python using the PyTorch ^52^ and MONAI ^45^ libraries on a Linux workstation with an AMD 5950X CPU, 32GB RAM, and an Nvidia RTX 3090 GPU. All data, code, and trained models used in this work have been made publicly available (see *Data Availability*, below).

## 3. Results

### 3.1 Synthetic datasets

All 14 parameter estimation methods were evaluated on all image series in the IMAGENET and URAND test datasets. An example image series from the IMAGENET test dataset is shown in Figure 3, with *T* maps estimated by all methods. In the high-SNR regions most methods produced similar-appearing results, although CNN_IMAGENET had distinctly lower noise. The estimated *T* maps were most different in the low-SNR regions, where the fitting methods showed high noise levels, whereas CNN_IMAGENET better recovered a noise-free *T* map, at the cost of modest blurring and distortion. Signed difference maps for each example in Figure 3 are provided in supplemental figure S1.

**Figure 3.**
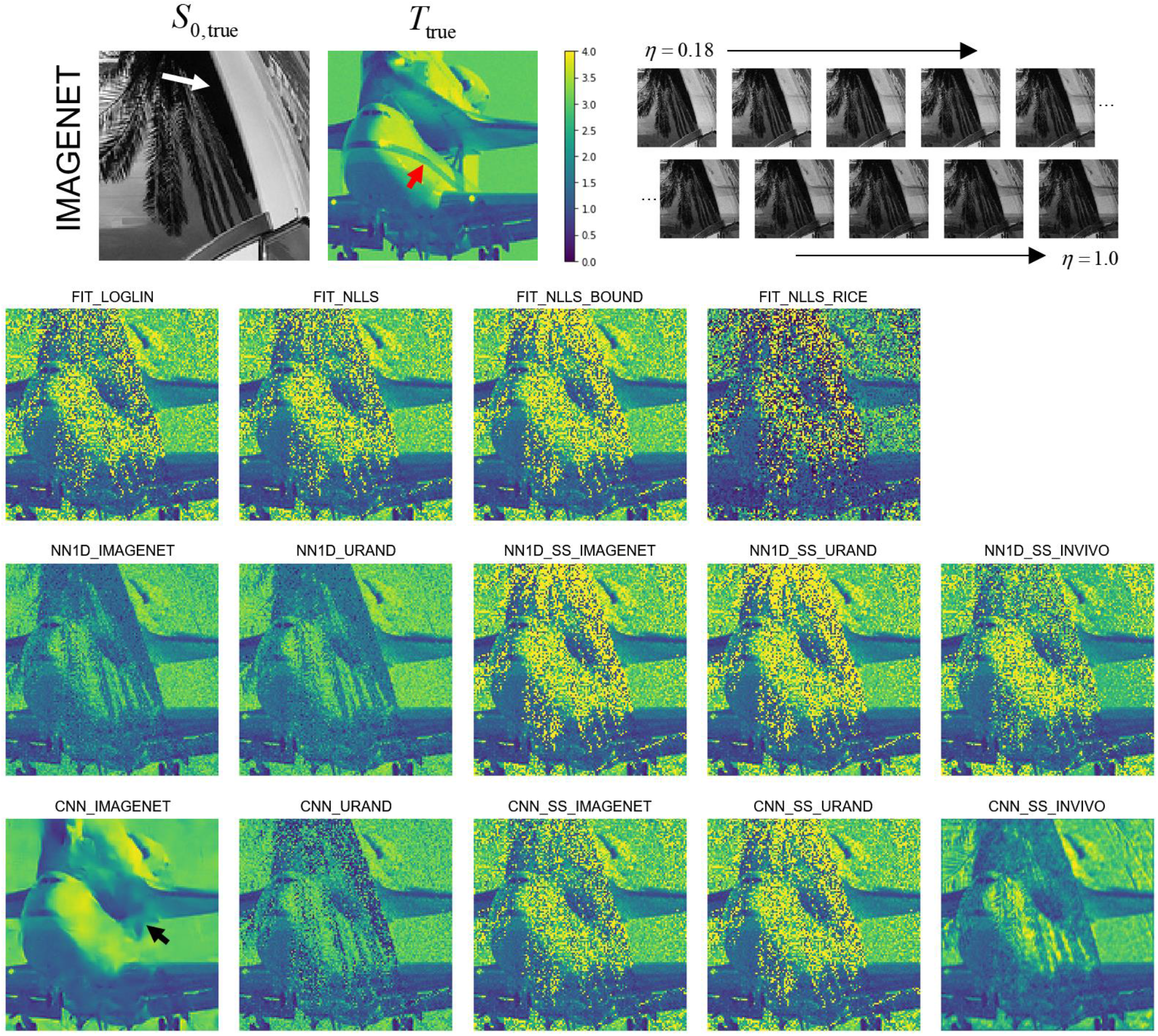
Example showing estimated *T* maps from all 14 methods for a case from the synthetic IMAGENET test dataset, with the true *S*_*0*_ and *T* map shown above. The regions of low *S*_*0*_ values (e.g., white arrow) have low SNR, which can be seen as regions of incorrect values in each of the estimated *T* maps. This example shows how the methods vary in performance at various noise levels and how they fail in regions of very low SNR. In this example CNN_IMAGENET had the smallest MAE (0.09) compared to the conventional FIT_NLLS (MAE=0.31) but shows evidence of blurring (black arrow) in the low SNR regions. Note also that none of the methods recovered the stripe visible in the *T*_true_ map (red arrow), underscoring the difficulty of this inverse problem.

Figure 4 provides a quantitative comparison of the methods’ performance in estimating *T* maps on both synthetic datasets. Focusing first on the four fitting methods, the two most common approaches (FIT_LOGLIN, FIT_NLLS) performed similarly, and exhibited a positive bias as expected. Incorporating bounds to the fitting (FIT_NLLS_BOUND) provided a small improvement of overall accuracy (i.e., reduced absolute error). The FIT_NLLS_RICE method had poorer accuracy and precision, likely due to the need to estimate three parameters instead of two. Considering the NN1D methods, those that were trained in a supervised manner (NN1D_IMAGENET and NN1D_URAND) had better overall accuracy than the self-supervised variants on both datasets. This is possibly because the self-supervised NN models do not fully model the Rician noise distribution, while this is implicitly learned with supervised training.

**Figure 4.**
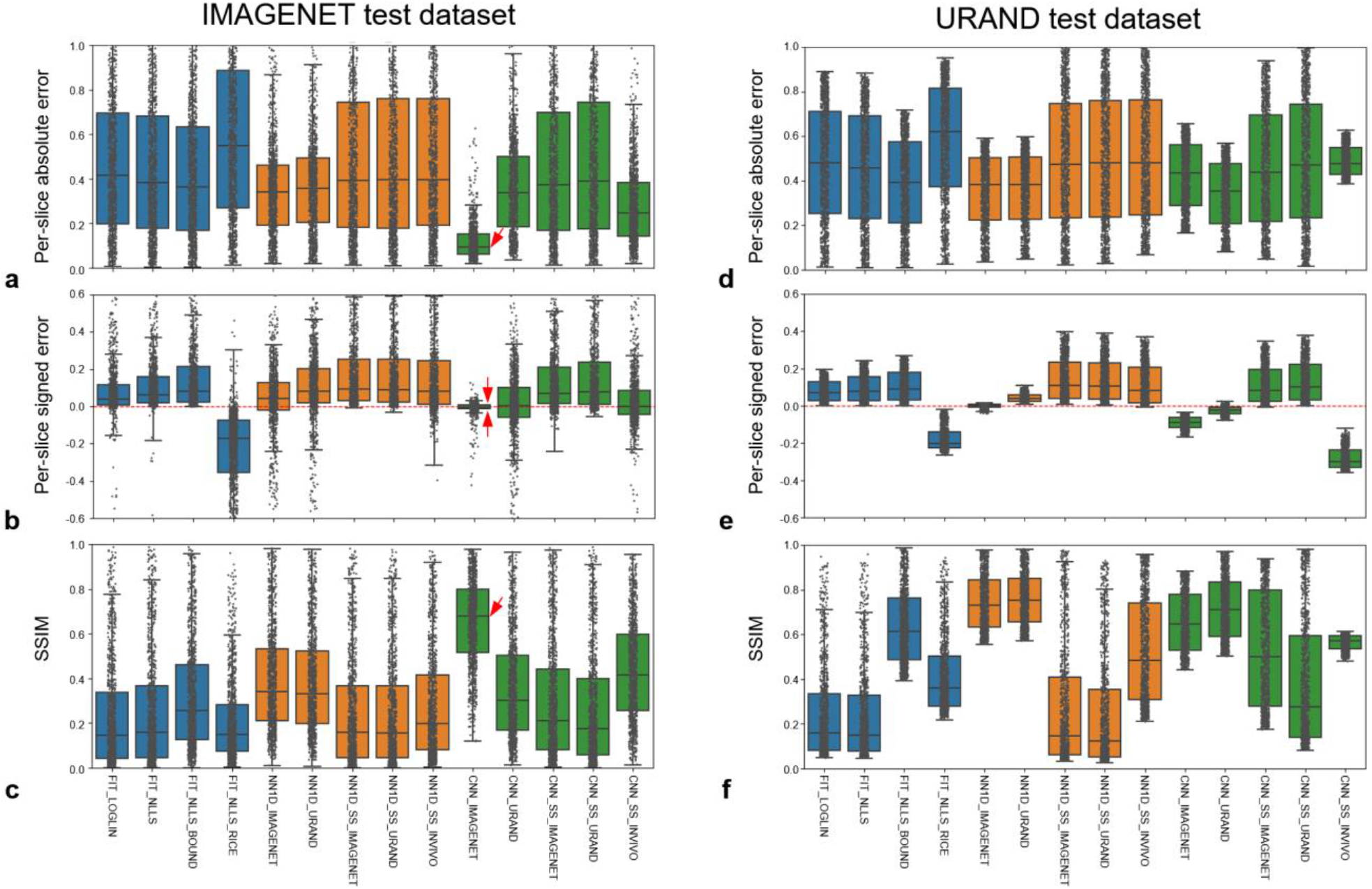
Comparative performance of the 14 methods for estimating *T* on the synthetic IMAGENET left column) and URAND (right column) test datasets, each consisting of 1000 image sets. The top row (a,d) plots the per-slice the absolute error (|*T*_pred_*-T*_true_|), the middle row (b, e) plots the signed error (*T*_pred_*-T*_true_), and the bottom row (c,f) plots the structural similarity between *T*_pred_ and *T*_true_. All box-whisker plots show median, interquartile ranges (IQR), and extrema (>1.5*IQR from quartiles), overlaid with the values from each of the 1000 cases. On the IMAGENET dataset, CNN_IMAGENET gave the lowest overall error (panel a), very low bias and highest precision (panel b), and the highest SSIM (panel c), as indicated by red arrows. Numerical values of this data are provided in supplemental table S1.

Looking at the CNN methods, the performance of CNN_IMAGENET on the IMAGENET test dataset stands out, showing low bias and the highest overall accuracy, precision, and structural similarity with the reference *T* maps. Notably, this method did not perform as well on the URAND test dataset. More generally, the fitting and NN1D methods performed similarly on the IMAGENET and URAND datasets, whereas CNNs gave higher accuracy on the IMAGENET dataset. This difference in performance suggests that, as expected, the CNNs use the information from spatially adjacent pixels to improve performance; when applied on data without spatial correlation the performance decreases.

For brevity, a subset of methods that performed well on the IMAGENET dataset of Figure 4 were selected for subsequent analyses: the conventional FIT_NLLS, the best performing CNN_IMAGENET, and the NN1D_URAND and CNN_SS_INVIVO methods as they both offered good performance and represent distinctly different methodologies.

Figure 5 presents the same experiment as Figure 4 but analyzed on a per-pixel basis rather than per-slice, in order to evaluate errors as a function of SNR and *T*_true_ values. The FIT_NLLS results (first column) demonstrate the tendency of this method to overestimate *T* when SNR is at low levels. CNN_SS_INVIVO (last column) had similar overall behavior. CNN_IMAGENET showed low variability and bias with relative consistency across values of SNR and *T*_true_, which are highly favorable characteristics for a *T* estimating method. NN1D_URAND had inconsistent behavior, both over- and underestimating *T* in different regimes of SNR and *T*_true_. Note that at high SNR, FIT_NLLS and CNN_IMAGENET gave similar performance, with low error and variability. An expansion of Figure 5 including all methods is provided in supplemental figure S2.

**Figure 5.**
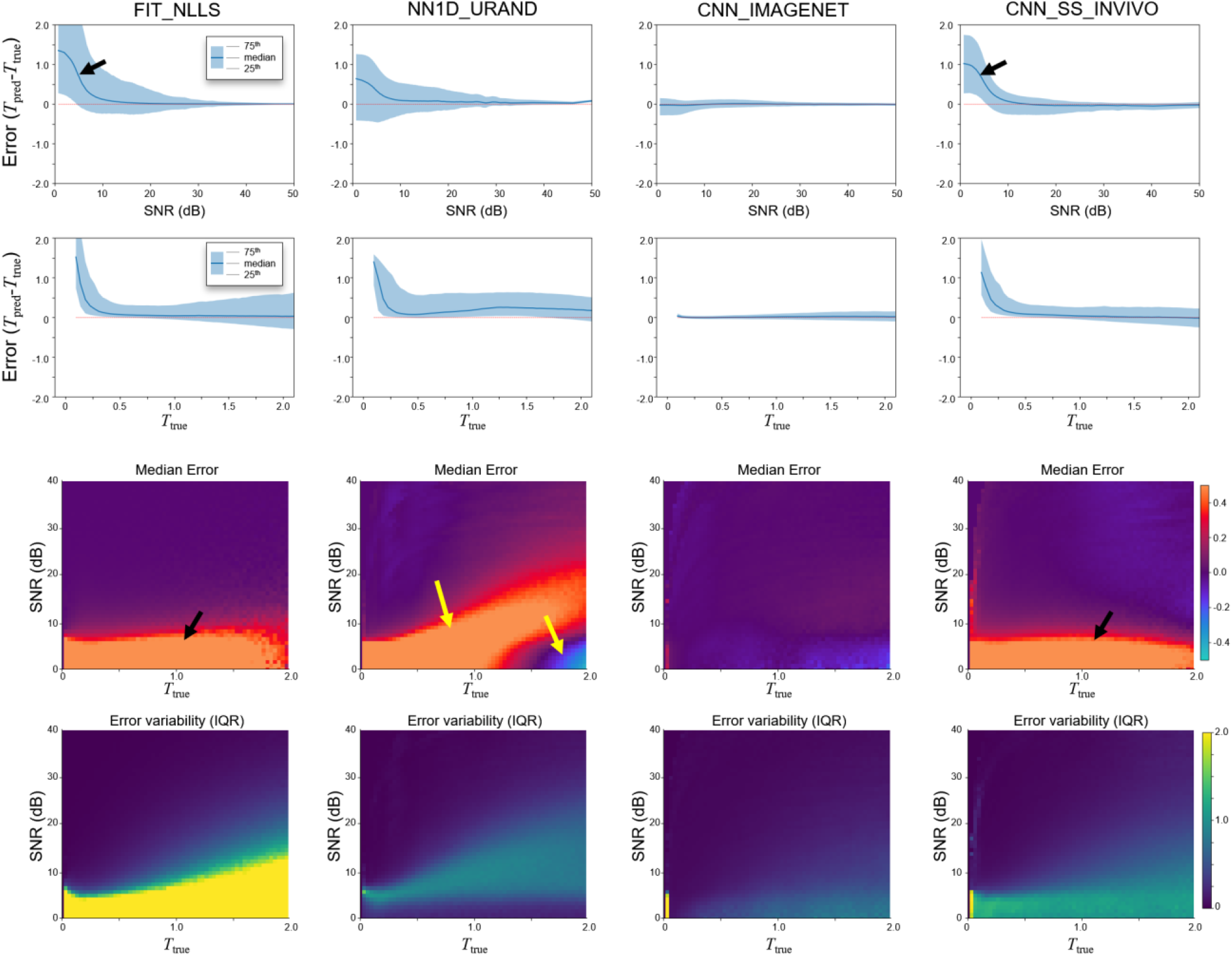
Estimation error (*T*_err_*=T*_pred_*-T*_true_) as a function of *T*_true_ and SNR for the four selected methods, on the IMAGENET test dataset, evaluated on a per-pixel basis. Both *T*_true_ and SNR dimensions were divided into 100 bins, and the median value and IQR (25^th^ and 75^th^ percentile) were calculated for each bin. Top row shows *T*_err_ as a function of SNR, while the second row shows the dependence on *T*_true_. The bottom two rows plot the median and IQR of *T*_err_ as a function of both *T*_true_ and SNR to show the interplay of these two effects. Both the FIT_NLLS and CNN_SS_INVIVO show positive bias at low SNR (black arrows), while CNN_IMAGENET has low bias and variability throughout. NN1D_URAND shows both over- and underestimation of T at low SNR (yellow arrow). Plots for all 14 methods are provided in supplemental figure S2.

### 3.2 In vivo dataset

An example comparing the four selected methods on an INVIVO test case is given in Figure 6. Qualitatively, the *T* maps for all methods appear generally similar, with a few exceptions: reconstruction artifacts can be seen in the zero-SNR rectum for both CNN_IMAGENET and CNN_SS_INVIVO, and the CNN_IMAGENET map shows less noise but with some evidence of blurring. Differences between the methods can be more clearly seen by separately focusing on regions with high (prostate, white arrow) and low (muscle, red arrow) SNR. CNN_IMAGENET gave similar values to FIT_NLLS in high-SNR regions, but lower values in low SNR regions. CNN_SS_INVIVO gave similar values to FIT_NLLS in both high- and low-SNR regions. NN1D_URAND gave variable values, with either higher or lower *T* values depending on SNR and *T*. All of these relationships are consistent with the results shown in the synthetic data. This suggests that the observations from the synthetic data apply to the *in vivo* case: CNN_IMAGENET gives accurate *T* estimates independently of *T* and SNR, whereas FIT_NLLS overestimates *T* in low SNR regions but gives accurate results where the SNR is high.

**Figure 6.**
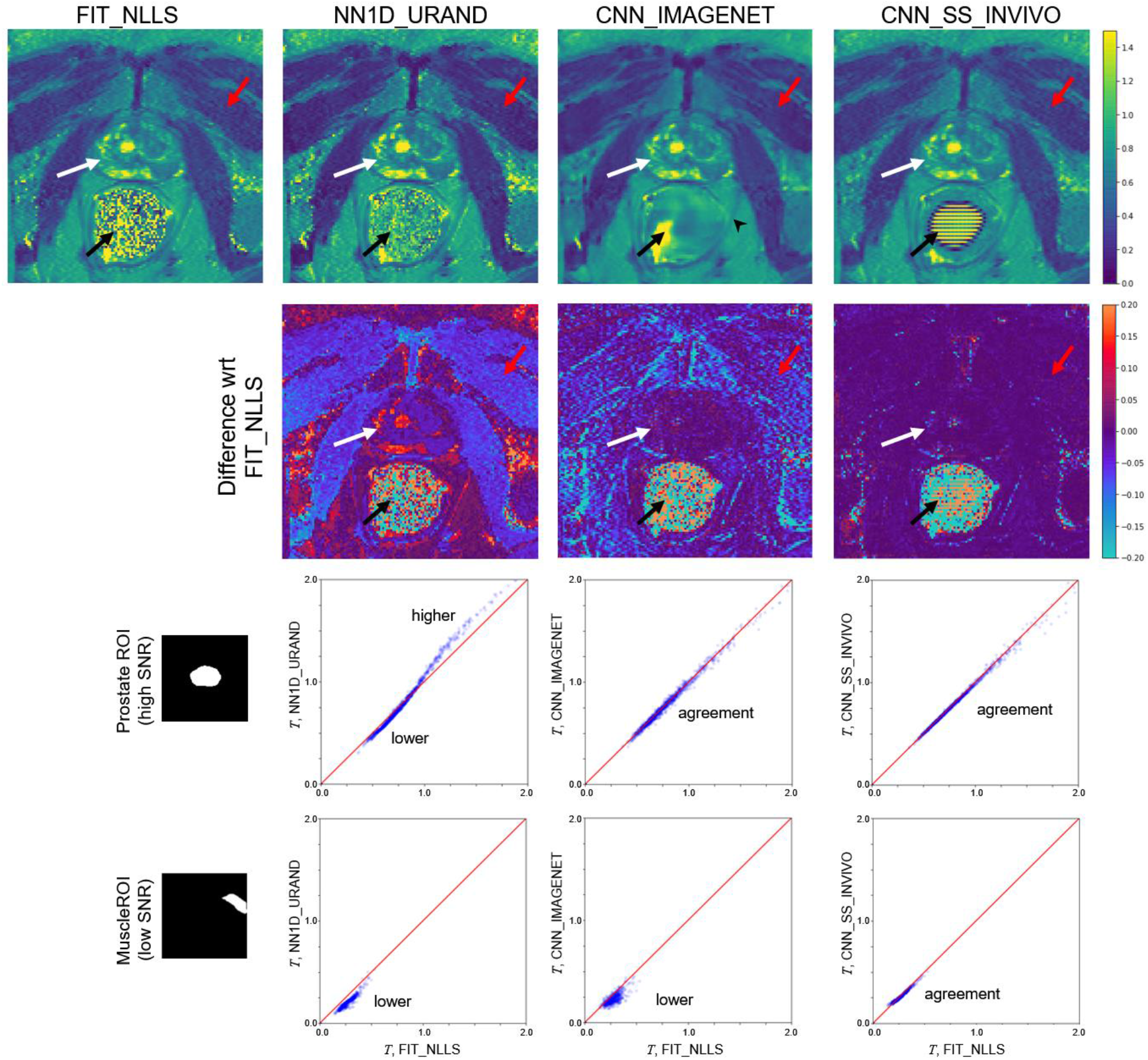
Comparison of selected methods on a single example slice from the INVIVO test set. Since no true value of *T* is available, the methods were compared to the standard FIT_NLLS. The top row shows *T* maps from each method; the second row shows the signed difference relative to FIT_NLLS. In the prostate (white arrow), where the SNR is high, CNN_IMAGENET and CNN_SS_INVIVO values were similar to FIT_NLLS, whereas NN1D_URAND gave *T* estimates that were more variable. In the low SNR muscle region (red arrow) results were less consistent, and in the rectum region (black arrow, where SNR is 0 due to a perfluorocarbon-filled balloon, CNN_IMAGENET and CNN_SS_INVIVO show reconstruction artifacts. The CNN_IMAGENET result appears least noisy, but shows blurring in some regions (e.g., black arrowhead). The bottom two rows show pixel-by-pixel comparisons of estimated *T* values over the ROIs in the prostate (high SNR) and a muscle region (low SNR). In the high SNR ROI, CNN_IMAGENET gives values similar to FIT_NLLS, but the low SNR ROI CNN_IMAGENET gives lower values, consistent the synthetic data.

The results of the noise addition experiment, shown in figures 7 and 8, also support this interpretation. Figure 7 shows an example image series from the INVIVO test dataset and the estimated *T* map from the three methods, with increasing amounts of retrospectively added Rician noise. Both the FIT_NLLS and CNN_SS_INVIVO methods showed increasing variation and increasing values for *T* at higher noise levels. In contrast the CNN_IMAGENET showed consistent values of *T*, but moderately increased blurring in the *T* maps. These trends can be seen across all cases in the INVIVO test dataset, as shown in figure 8. The CNN_SS_INVIVO model performed similarly to FIT_NLLS, but the CNN_IMAGENET model showed greater robustness to noise, with smaller changes in accuracy, bias, precision, and SSIM relative to the *T* maps calculated from the original (no noise added) data.

**Figure 7.**
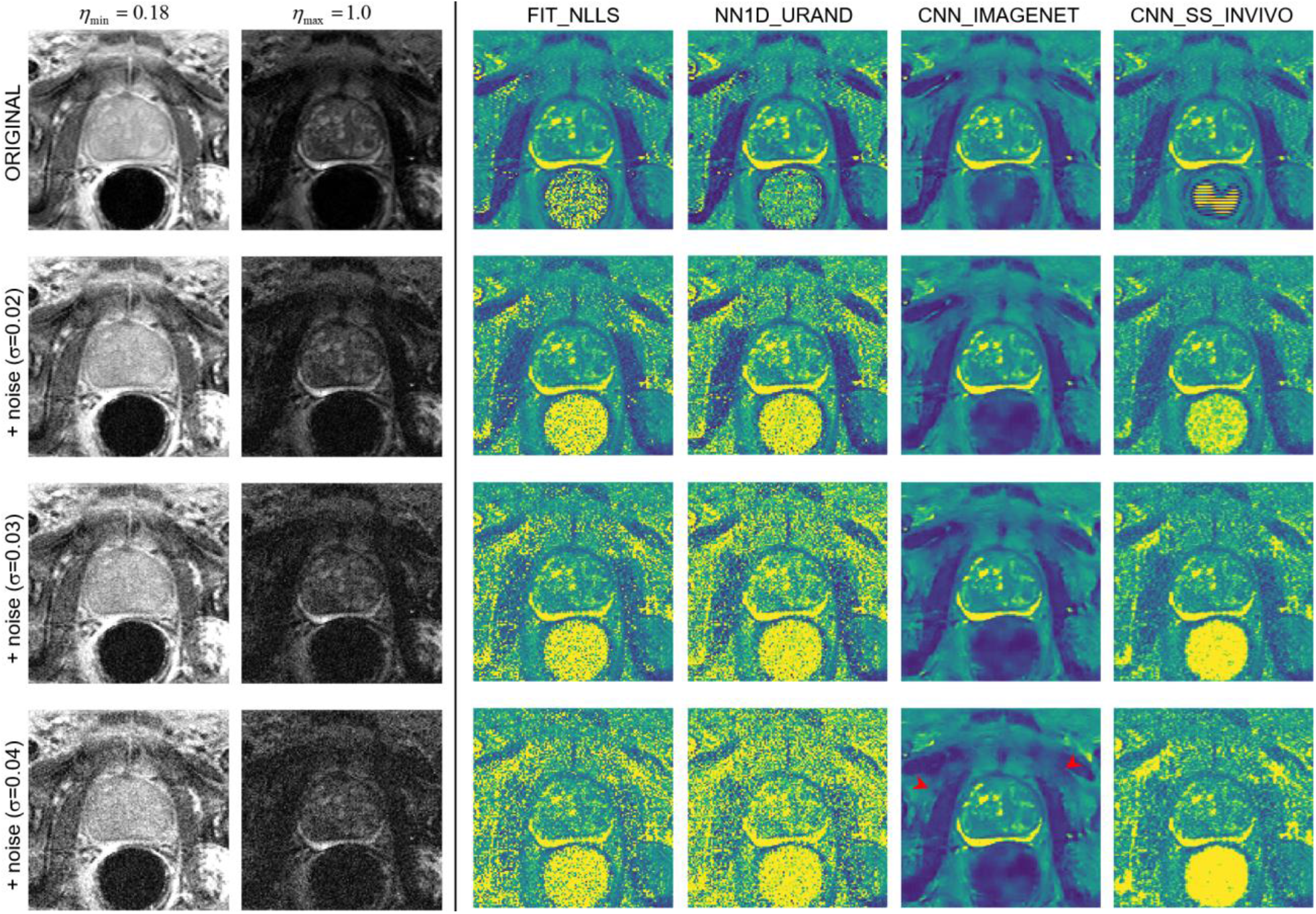
An example case from the noise-addition experiment. The top row shows the unmodified data: images of the shortest and longest echo time, and *T* maps calculated with the four selected estimation methods. The next three rows show the same data with increasing noise added (with Gaussian standard deviation 0.02, 0.03, and 0.04), and the corresponding *T* maps. At higher levels of added noise, the predicted *T* maps from FIT_NLLS and NN1D_URAND show increasing noise and increased higher values of *T* throughout the image. CNN_SS_INVIVO shows similar behavior, with moderately less noise and some evidence of blurring. In contrast, CNN_IMAGENET shows modest blurring (red arrowheads) and no noise amplification with increasing added noise.

**Figure 8.**
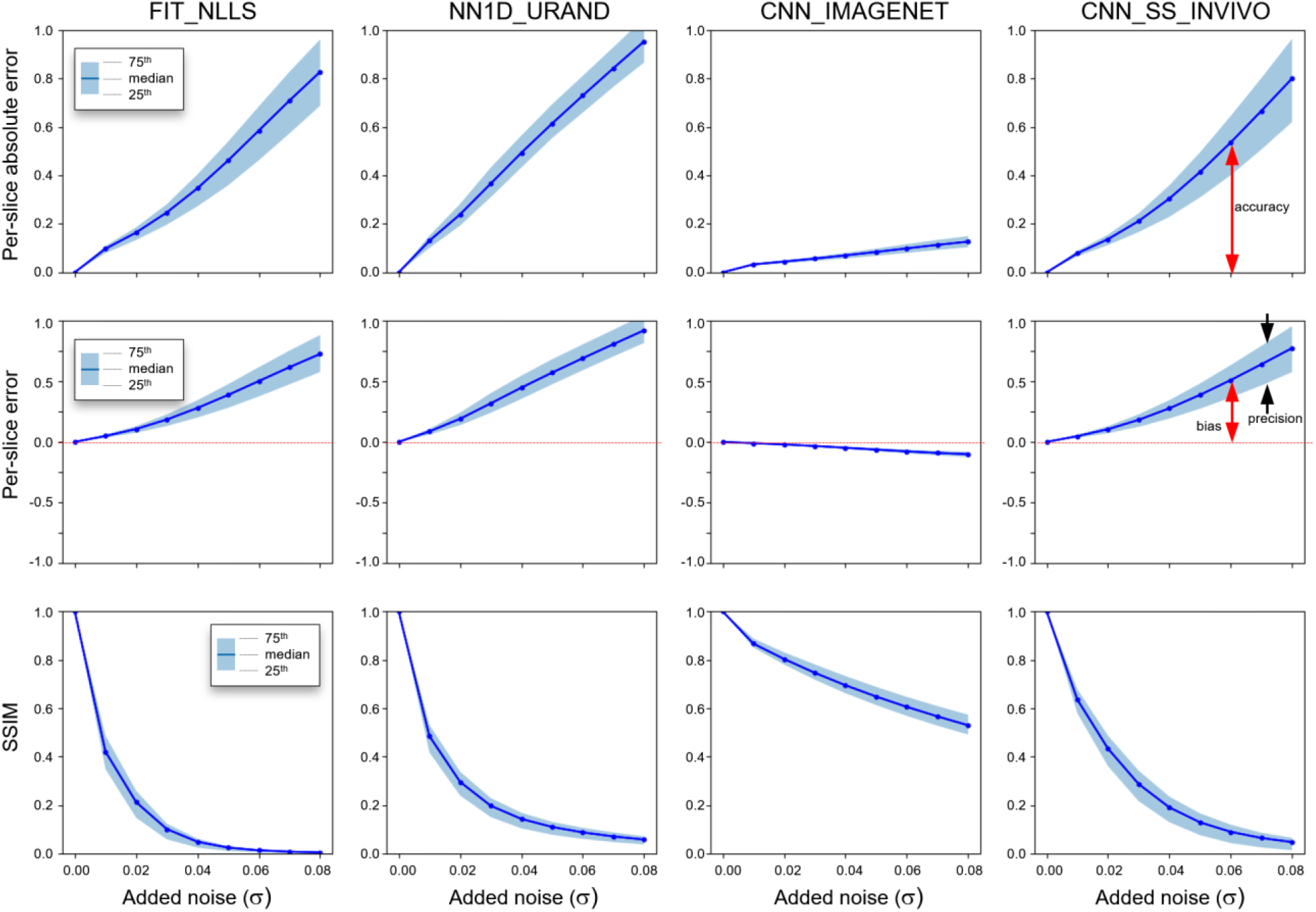
Results from the noise-addition experiment showing the per-slice median absolute error (top row), per-slice signed error (middle row), and SSIM relative to the reference *T* map, which is the map calculated using the same method on the original (no noise added) dataset. Plots show the median and IQR of all 694 slices in the INVIVO test set as the level of added noise increases from left to right. Annotations indicating definitions of bias, precision, and accuracy are included for clarity. CNN_IMAGENET shows the least change in these three metrics with increasing added noise.

## 4. Discussion

In this work we compared a large variety of methods for estimating T_2_ from prostate T_2_ relaxometry acquisitions. Our main finding is that a CNN, trained in a supervised fashion with a physics-based synthetic dataset (CNN_IMAGENET), gave the best overall accuracy when tested on simulated data. Additionally, when evaluated on *in vivo* data the performance was similar to that of the simulated evaluation, giving values in agreement with conventional NLLS fitting in the high SNR regime, and providing better precision and a likely reduction in bias in low SNR regions.

Importantly, the comparison amongst the ten NN variations and four fitting techniques provides insight into three critical factors that enable CNN_IMAGENET to outperform the conventional NLLS approach. The first factor is the ability of the synthetic training strategy to correctly incorporate the Rician noise distribution. While the problem of least-squares fitting in Rician noise is widely recognized ^23,30^, the solutions proposed require knowledge of the noise distribution on a per-pixel basis, which is not easily determined retrospectively from images reconstructed with parallel imaging. Parallel imaging is routinely used to reduce acquisition times but produces images with a spatially-varying noise distribution. Trying to estimate the pixel-wise noise simultaneously with the relaxation rate increases the degrees of fitting of the model and leads to additional error and bias in the other parameter estimates.

The Rician noise distribution affects not just the NLLS methods but also the self-supervised NN models. These were trained with a least-squares loss on data with Rician noise, and consequently they show a positive bias for *T* at low SNR, just like the NLLS methods. It may be possible to design training loss functions that incorporate the Rician distribution. We were not successful in training such NNs due to the numerical instability of the Bessel functions that analytically describe the distribution ^25,30,43,53^, but this may be possible using approximate loss functions or different training methods.

The Rician noise issue is a consequence of the specific problem domain we chose to focus on: fitting previously reconstructed magnitude images with spatially varying noise. This issue can be avoided by fitting real- or complex-valued data ^54,55^, or by generating accurate noise maps from calibration acquisitions, but these are not generally output from MR scanners, and can be sensitive to phase errors. Our focus has broad practical value: it allows these methods to be used for retrospective analyses of conventionally-acquired relaxometry datasets available in DICOM format, and it can be more readily applied to clinical studies without needing custom pulse sequences and reconstructions.

The second factor contributing to improved quantitative performance is that NNs trained in a supervised fashion produce outputs that are limited by the range of values present in the training data. An unconstrained NLLS fit can lead to a very large range of parameter estimates, particularly in noisy data. Using a constrained fit (FIT_NLLS_BOUND) with relatively wide limits substantially reduces the error compared to unbound fitting. The synthetic datasets used for training had values of T drawn from a uniform random distribution over the same range as the bounds in FIT_NLLS_BOUND (*T* ϵ[0.045, 4]), so these methods had similar range constraints.

The distribution of parameters in the synthetic training data limits the range of values that are produced by NN inference, but they can also bias the output values. This bias can be undesirable ^56^, but the negative effects can be avoided by careful selection of the training range. In T_2_ mapping, the TE array is generally selected to cover the range of T_2_s the investigators anticipate. The bias of expected T_2_ values is essentially built directly into the acquisition parameters. By synthesizing training data with uniform distribution of T_2_ values, over a wide range (from 0.25 times the shortest TE value to 4 times the largest), the negative impact of bias is minimized.

The third factor is the convolution operator, which takes advantage of the spatial correlation between pixels and improves performance in low SNR regions. The training strategy used for CNN_IMAGENET, in which the network attempts to predict noise-free *T* maps from noisy image series, provides inherent denoising, as the model implicitly “learns” the Rician distribution and seeks to remove it. The spatial convolution operator is a key component of this denoising process – we showed that a 1D network trained on the same data (NN1D_IMAGENET) exhibited lower overall precision (Figure 4) and greater estimation variance at low SNR levels (see supplemental figure S1).

Improvement in noise robustness comes at the cost of some blurring, which can be seen in our data as well as in prior studies ^37^. The amount of blurring depends on the architecture, training strategy, and loss functions. Zhao et al. ^57^ attribute this phenomenon to the use of an L2 loss function, and have proposed alternate loss functions that could improve the perceived image quality. Improving these artifacts, while maintaining quantitative performance, is a topic for future work.

The strategy of synthetic supervised training, using a large synthetic dataset derived from a physics-based signal model, has broad applications in the MR field. As first demonstrated with the AUTOMAP image reconstruction method ^41^, this approach can be used to build large datasets for training, and can encode signal models with much greater complexity than the simple monoexponential decay shown here. The dataset synthesis encodes the forward signal model, and through the training process the network learns a mapping that encodes the inverse signal model. In this work we demonstrated its application in prostate T_2_ relaxometry, but this same strategy can be used for other relaxometry applications, diffusion modeling, and potentially problems with more complex signal models.

In addition to improving retrospective analyses, the methods demonstrated herein could be used to optimize acquisitions for prospective studies. With increased robustness to low image SNR, it may be possible to acquire data with higher accelerations or spatial resolution without losing quantitative performance. Furthermore, since the CNN_IMAGENET method has lower variability than FIT_NLLS in estimating T_2_ at long values (e.g., T_2_ > TE_max_, or *T* >1, as shown in figure 5), it may be possible to acquire shorter echo trains to reduce heating (specific absorption ratio) and/or increase the number of slices acquired within the same scan time.

This study has several limitations. This work used relatively simple NN architectures and training strategies, which could be improved upon using architectural variations, different loss functions, and expanded or augmented training datasets. A disadvantage of the model architectures used is that they are sized and trained to work only for a specific set of *η* values; to apply this approach to data with different TE values, one would need to generate a new synthetic dataset and train a new model for that specific set of parameters. We also did not compare our methods with Bayesian fitting ^58,59^ or dictionary-based parameter estimation, two additional strategies that merit further exploration. These methods could incorporate Rician noise and restricted parameter ranges but would not easily incorporate the learning of spatial priors provided by CNNs. Finally, we did not evaluate the performance on *in vivo* data from multiple sites, vendors, or field strengths. Since no information about these factors was used in generating the synthetic training data, the performance may extend to heterogeneous datasets, but this assessment was not performed.

## 5. Conclusions

We compared conventional NLLS fitting with several neural network architectures and training strategies for estimating T_2_ maps from multi-echo magnitude images acquired for prostate relaxometry. We found that a CNN, trained with synthetic data in a supervised manner, gave better accuracy and noise robustness than NLLS fitting and other NN methods. By comparing the performance of different estimation methods on multiple synthetic datasets we were able to identify three specific factors that led to the performance gains: improved Rician noise modeling, restriction of the parameter estimation domain, and learning of spatial priors with convolutional layers. Furthermore, we showed the feasibility of using these CNNs for analyzing *in vivo* prostate T_2_ relaxometry data and demonstrated its excellent performance in the low SNR regime.

## Data Availability

All data produced are available online at the UMN Data Conservancy. All code and trained models are available on Github.

https://conservancy.umn.edu/handle/11299/243192

https://github.com/patbolan/prostate_t2map

## Abbreviations

CNN: convolutional neural network
IQR: interquartile range
NLLS: non-linear least squares
NN: neural network
SNR: signal-to-noise ratio
SSIM: structural similarity index measure

## Acknowledgements

Funding sources NIH P41 EB027061, NIH R01 CA241159, NIH R01 EB029985, NIH S10 OD017974-01, NIH R01 HL153146, NIH R21 EB028369

## Data Availability

All of the de-identified *in vivo* data used in this study are available in the Data Repository for the University of Minnesota, at https://conservancy.umn.edu/handle/11299/243192. The source code for training neural networks, evaluating results, and generating the figures in this manuscript, along with the trained models, are available on Github at https://github.com/patbolan/prostate_t2map.

## Supplemental Data

**Figure S1.**
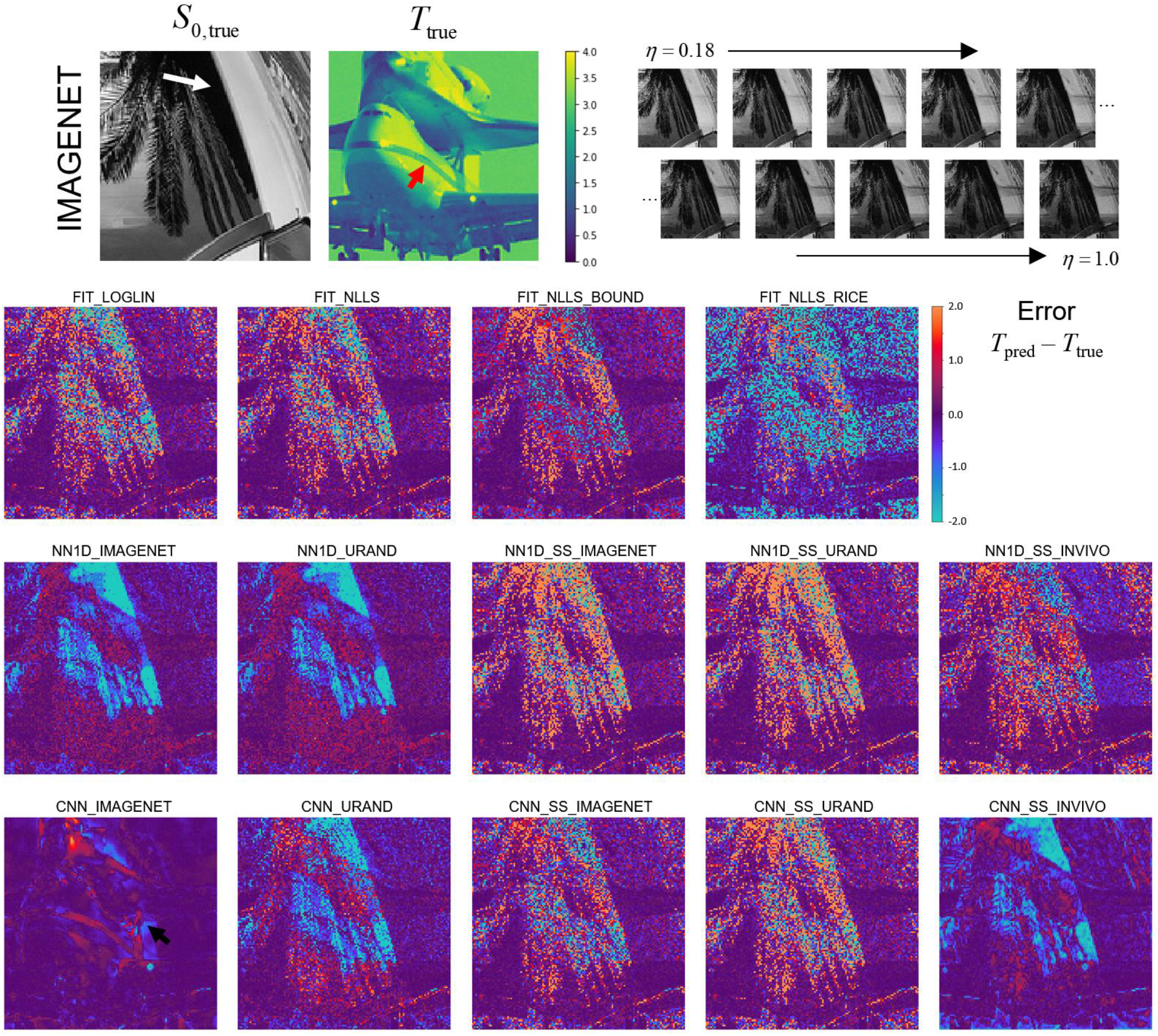
Image maps showing the signed error between predicted and true *T* maps for each of the methods, using the example case provided in figure 3. Red pixels indicate overestimation of *T* (positive errors) while blue pixels indicate underestimation. All values in normalized *T* units.

**Figure S2:**
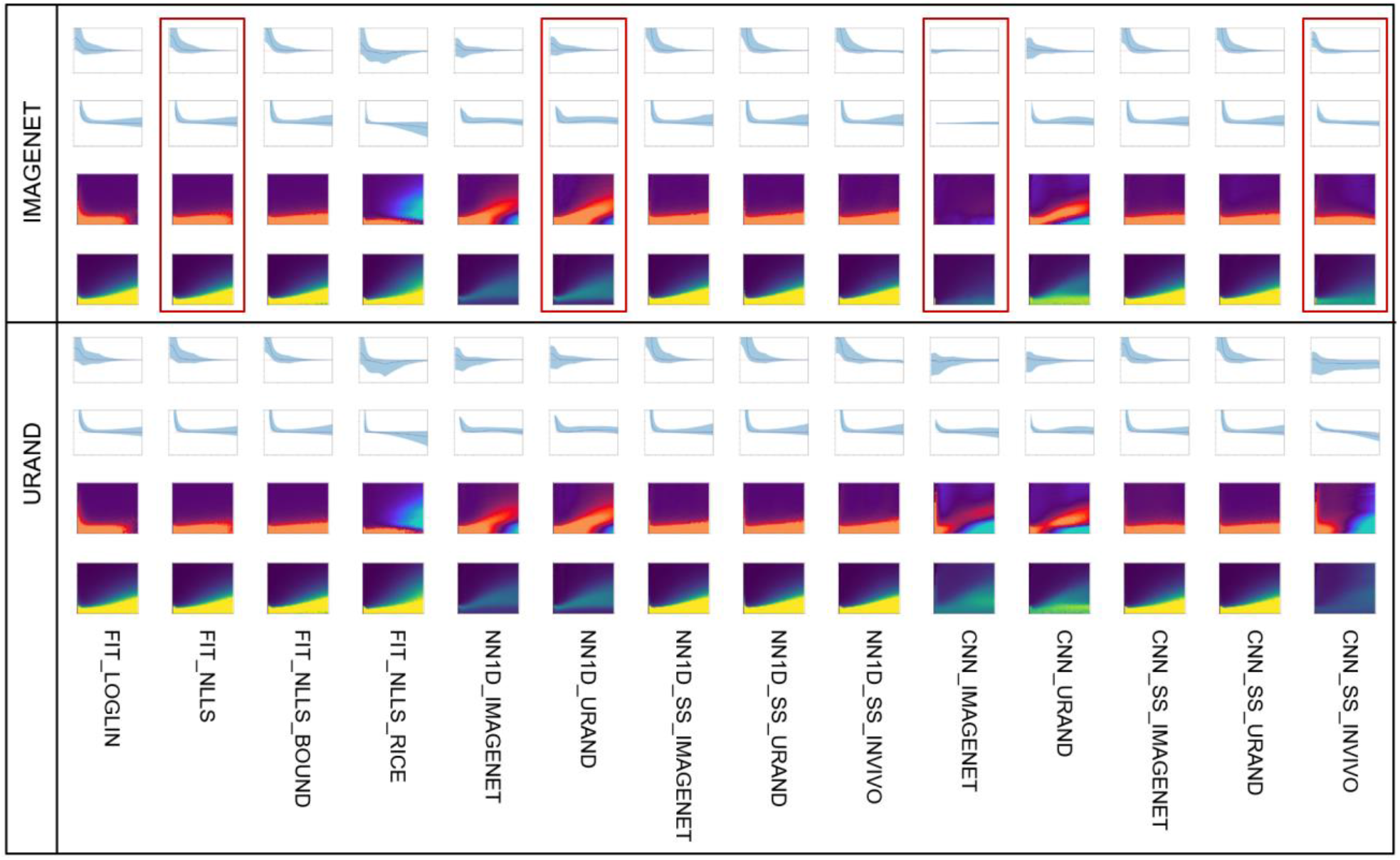
Expansion of figure 5, showing *T*-estimation error (*T*_err_=*T*_pred_-*T*_true_) with all 14 methods on both synthetic datasets. The four cases shown in Figure 4 are outlined in red. An ideal estimator of *T* would be deep blue (uniformly zero error) in the colormaps, independent of both SNR and *T*_true_; CNN_IMAGENET best approximates this ideal method when evaluated on the IMAGENET test dataset. See Figure 4 for further explanation and axis labeling.

**Table S1.** Numerical values of the primary metrics plotted in Figure 4. The best values for each dataset (lowest bias, precision, and overall error; highest SSIM) are shown in **bold**.

| Dataset | Method | Bias | Precision | Error | SSIM |
| --- | --- | --- | --- | --- | --- |
| IMAGENET_TEST_1K | FIT LOGLIN | 0.038 | 0.109 | 0.416 | 0.146 |
|  | FIT NLLS | 0.063 | 0.143 | 0.383 | 0.160 |
|  | FIT NLLS BOUND | 0.081 | 0.191 | 0.363 | 0.257 |
|  | FIT NLLS RICE | -0.173 | 0.282 | 0.549 | 0.149 |
|  | NN1D IMAGENET | 0.042 | 0.149 | 0.340 | 0.342 |
|  | NN1D URAND | 0.083 | 0.182 | 0.358 | 0.330 |
|  | NN1D SS IMAGENET | 0.094 | 0.224 | 0.394 | 0.159 |
|  | NN1D SS URAND | 0.089 | 0.229 | 0.395 | 0.157 |
|  | NN1D SS INVIVO | 0.082 | 0.233 | 0.397 | 0.197 |
|  | CNN IMAGENET | **-0.002** | **0.017** | **0.093** | **0.678** |
|  | CNN URAND | **0.002** | 0.157 | 0.336 | 0.303 |
|  | CNN SS IMAGENET | 0.069 | 0.198 | 0.374 | 0.211 |
|  | CNN SS URAND | 0.078 | 0.228 | 0.391 | 0.176 |
|  | CNN SS INVIVO | **-0.002** | 0.127 | 0.248 | 0.415 |
| URAND_TEST_1K | FIT LOGLIN | 0.071 | 0.100 | 0.480 | 0.158 |
|  | FIT NLLS | 0.079 | 0.130 | 0.457 | 0.149 |
|  | FIT NLLS BOUND | 0.090 | 0.147 | 0.393 | 0.613 |
|  | FIT NLLS RICE | -0.203 | 0.085 | 0.621 | 0.359 |
|  | NN1D IMAGENET | **0.001** | **0.016** | 0.383 | 0.731 |
|  | NN1D URAND | 0.038 | 0.037 | 0.383 | **0.752** |
|  | NN1D SS IMAGENET | 0.111 | 0.196 | 0.475 | 0.146 |
|  | NN1D SS URAND | 0.105 | 0.196 | 0.480 | 0.124 |
|  | NN1D SS INVIVO | 0.084 | 0.191 | 0.480 | 0.483 |
|  | CNN IMAGENET | -0.089 | 0.055 | 0.434 | 0.647 |
|  | CNN URAND | -0.023 | 0.036 | **0.355** | 0.711 |
|  | CNN SS IMAGENET | 0.083 | 0.173 | 0.437 | 0.499 |
|  | CNN SS URAND | 0.100 | 0.189 | 0.469 | 0.275 |
|  | CNN SS INVIVO | -0.301 | 0.097 | 0.479 | 0.571 |

